# Natural history and epidemiology of cleft lip and palate: a registry-based study in Iran (2000-19)

**DOI:** 10.1101/2022.01.02.22268601

**Authors:** Mahdi Biabani, Saeed Dastgiri, Elham Davtalab Esmaeili

**Affiliations:** School of Dentistry, Tabriz University of Medical Sciences, Tabriz, Iran; Tabriz Health Services Management Research Center, Tabriz University of Medical Sciences, Tabriz, Iran; Road Traffic Injury Research Center, Tabriz University of Medical Sciences, Tabriz, Iran

**Keywords:** Cleft lip, Cleft palate, Epidemiology, Prevalence, Mortality, Survival

## Abstract

The aim of this study was to provide the natural history and epidemiology of cleft lip and cleft palate in the northwest region of Iran between 2000 and 2019. Since 2000, infants born with birth defects have been registered in the Tabriz Registry of Congenital Anomalies (TRoCA). For this study, the information and data were collected using the TRoCA registry system. Prevalence of cleft lip and cleft palate was 1.48 (95% CI 1.34; 1.62) per 1000 live births over the past two decades in the region. The occurrence of cleft lip and cleft palate was more common in males than females. The fetal death ratio was 5 percent of live born children. The proportion of infants with cleft lip and cleft palate surviving to the second week was 54 percent. The results may have a role in planning and evaluating the strategies for primary prevention of cleft lip and cleft palate, particularly in high-risk populations.

## Introduction

Cleft lip and palate is the most common major congenital anomalies affecting the orofacial area. It is the second most common defect in newborns in many places (1-4). Research reports indicate that both genetic and environmental factors contribute to the occurrence of these defects (5, 6).

Many studies reported different figures for the prevalence of cleft lip and cleft palate in different geographical areas of the world. Accordingly, Indian American communities showed the highest, and blacks the lowest occurrence rate of defect. This rate was reported higher in some of the Asian countries too (7). Prevalence rate of these anomalies was 1 (per 1000 births) in Iran. This is lower than other countries in the region (8).

Infants born with cleft lip and cleft palate have many problems including dental deformities and malocclusion as well as difficulty in chewing, swallowing, speaking and esthetic requiring rehabilitation and a team of specialists to repair (9). Assessing the prevalence of cleft lip and palate would be helpful in planning and providing health care services for these children, and the information obtained can also be used as a source for future researches (10). The main aim of this registry-based study was to provide the natural history and epidemiology of cleft lip and cleft palate in the northwest of Iran between 2000 and 2019.

## Materials and Methods

Since 2000, infants born with birth defects in the region have been registered in the Tabriz Registry of Congenital Anomalies (TRoCA). The registry continues to operate under the guidelines of the World Health Organization (WHO), and as a member of the International Clearinghouse for Birth Defects Surveillance and Research (ICBDSR).

For this study, the information and data for cleft lip and palate including gender, birth weight, maternal age at delivery, ABO and Rh blood groups, social status, and the presence of concomitant diseases and abnormalities were collected using the TRoCA registry system. The International Classification of Disease (ICD -10) was used for coding the congenital anomalies. For the purpose of data analysis, prevalence rates with 95 percent confidence intervals were calculated.

This study has received ethical approval from Tabriz University of Medical Science (IR.TBZMED.REC.1399.624).

## Results

A total of 291,569 live births were investigated by the TRoCA registry from which 431 cases were identified as having cleft lip and cleft palate presenting a total prevalence rate of 1.48 (95% CI 1.34; 1.62) per 1000 live births over the past two decades in the region. The prevalence rate of these anomalies during the first 10 years (2000-09) and the second 10 years (2010-19) estimated 1.05 (95% CI 0.91; 1.19) and 2.46 (95% CI 2.13; 2.78) per 1000 live births. The prevalence rate of cleft lip with/without cleft palate was 0.94 (95% CI 0.82; 1.05) and the prevalence rate of isolated cleft palate was 0.54 (95% CI 0.45; 0.62) per 1000 live births. Figure 1 and 2 shows the prevalence rate and the time trend of cleft lip with/without cleft palate and isolated cleft palate in the area during the study period.

**Fig 1.**
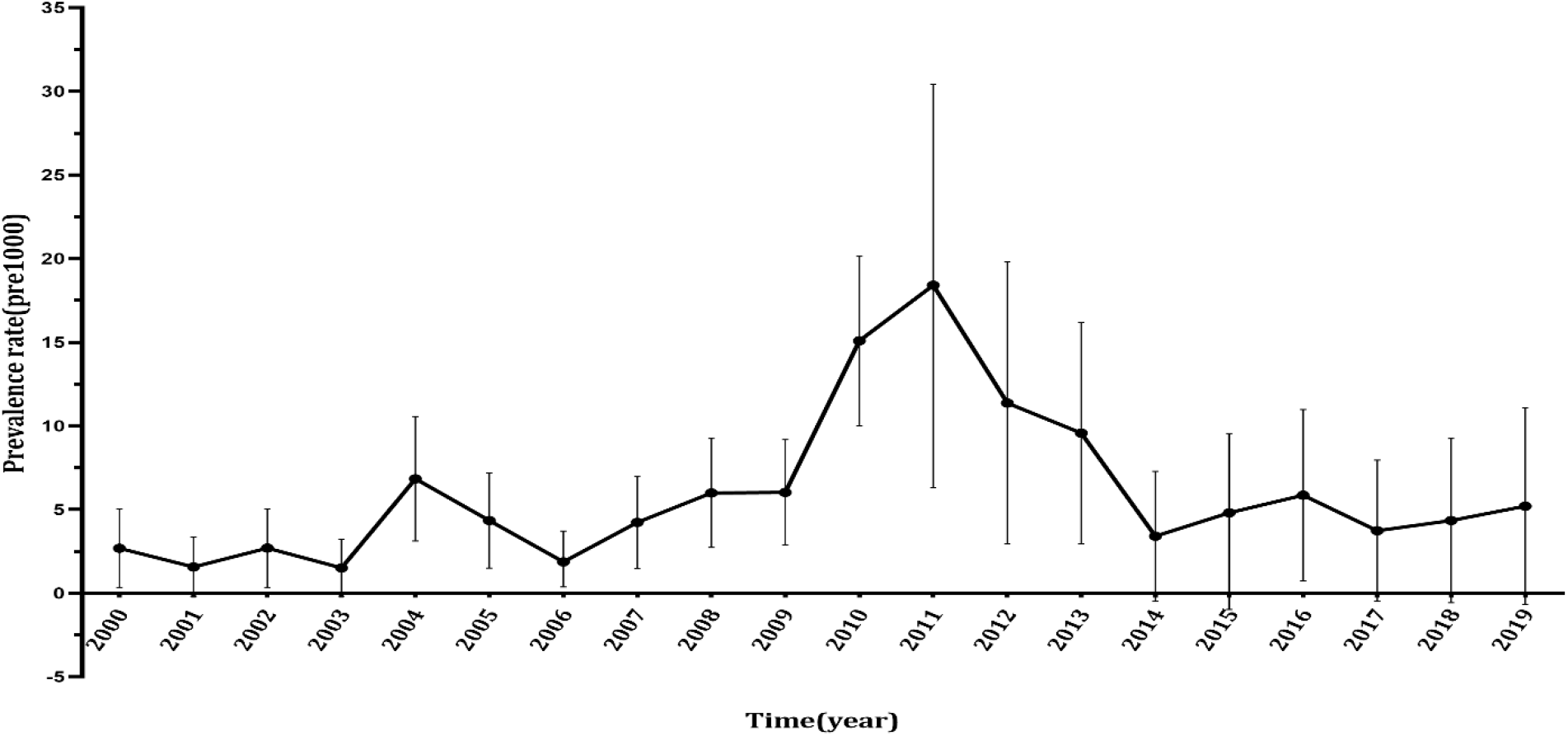
The prevalence (95 percent CI) and the time trend of isolated cleft palate in 1000 live births in the northwest of Iran (2000-19)

**Fig 2.**
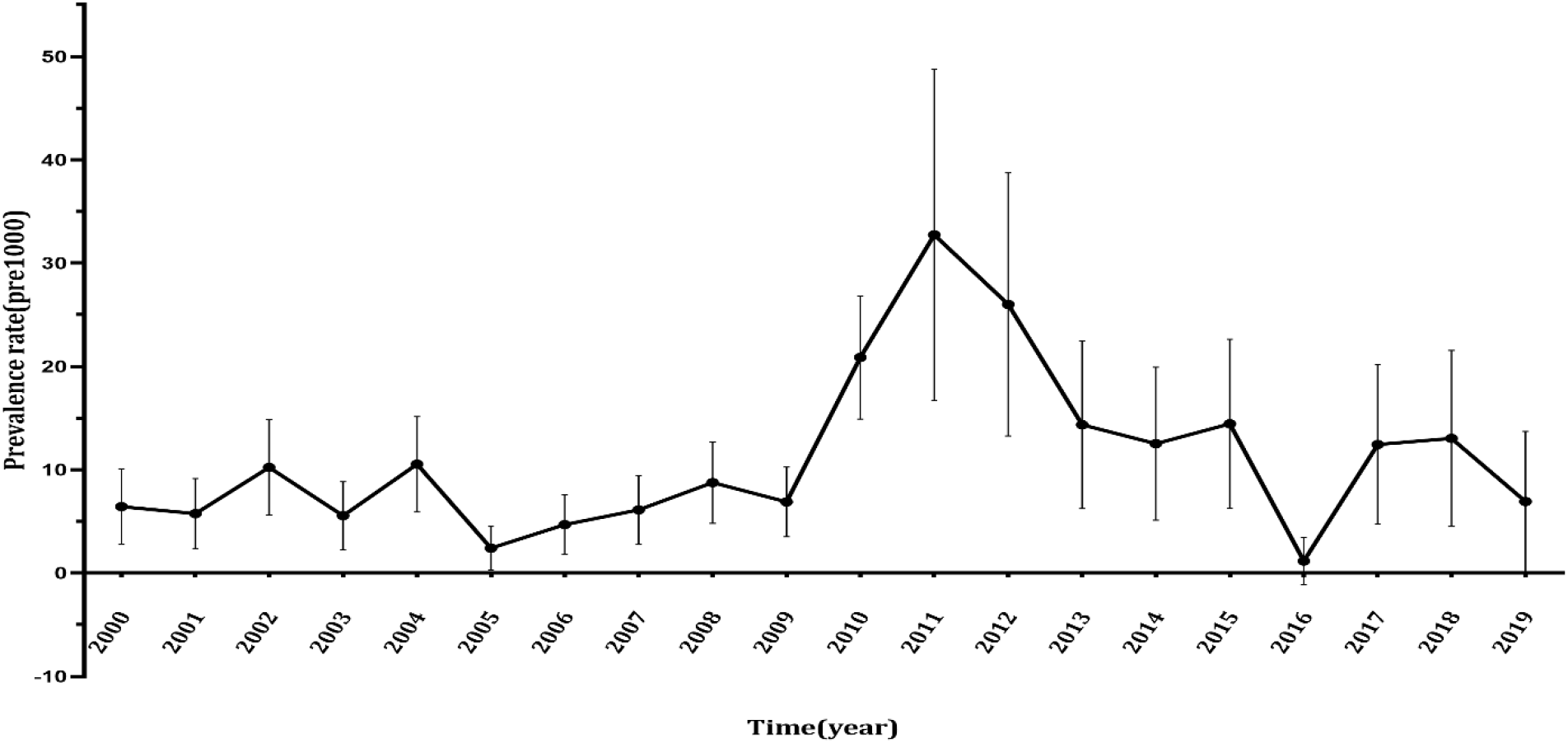
The prevalence (95 percent CI) and the time trend of cleft lip with/without cleft palate in the northwest of Iran (2000-19)

The occurrence of cleft lip and cleft palate was more common in males than females while the prevalence of isolated cases was higher in females. The occurrence of these defects by newborn weight, maternal age at delivery, social status, and maternal blood type are shown in table 1.

**Table 1.**
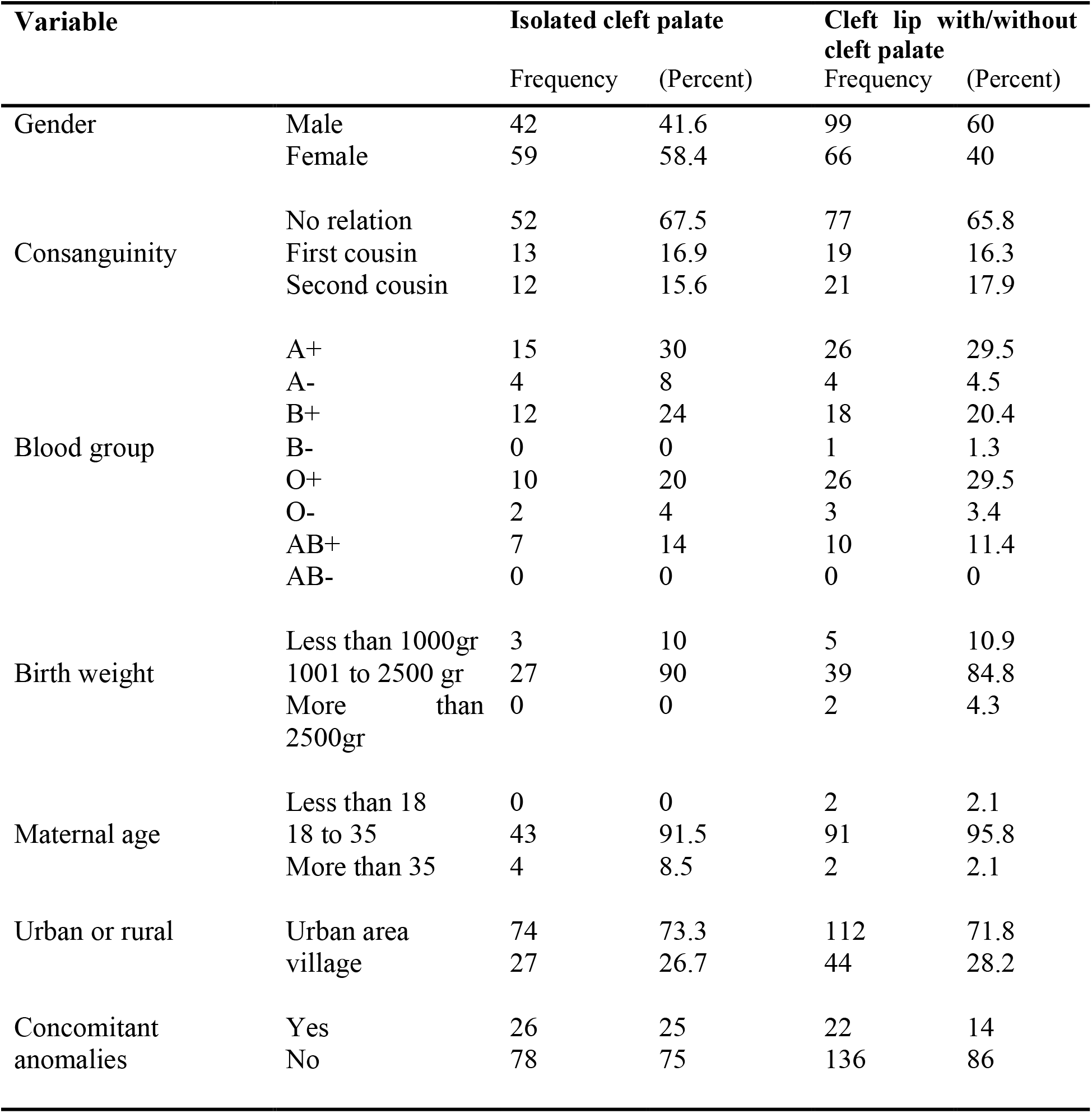
Characteristics of the infant with cleft lip and cleft palate in the northwest of Iran (2000-19)

A total of 52 (67.5 %) and 77 (65.5 %) of parents of isolated cases with cleft palate and cleft lip with/without cleft palate had no consanguineous marriage, respectively. The A+ and O+ blood groups were more common types among cleft lip with/without cleft palate. Type A+ was reported in 15 (30 %) of isolated cleft palate. Birth weight of 1000-2500 grams observed in 27 (90 %) and 39 (84.8 %) of isolated cleft palate and cleft lip with/without cleft palate cases, respectively. A number of 43 (91.5 %) of isolated cleft palate and 91 (95.8 %) of cleft lip with/without cleft palate born from mother aged 18 to 35.

Fetal death ratio was 5 percent of live births. Mortality rate of these groups of anomalies was 5 percent among all birth defects. The proportion of infants with cleft palate and cleft lip surviving to the second week of life was 54 percent.

## Discussion

In this study, the total prevalence rate of cleft lip and cleft palate showed an increasing trend over the past two decades in the region. This rate is two times more than the rate reported from Iran by Khazaei et al (8). Although similar studies conducted in different region of Iran, reported prevalence rates of these anomalies lower than of the current investigation, Jalili et al in the study from the capital city, Tehran reported similar figure with us (11-15).

Saudi Arabia (0.3 per 1000 live births) and Jordan (1.4 per 1000 live births) have a lower prevalence rate than ours whereas Pakistani rate was higher with 1.9 per 1000 live births (16-18). Beside similar figures reported in many other studies, Greg et al and Fitzpatrick et al reported different rates (19-23). Environmental factors, ethnic variations, maternal nutrition, and consanguineous marriages might have a role in explaining these differences.

Most of the infants born with these anomalies were in the weight range of 1000-2500 grams. We had no control group to compare, however, Lei et al found in Taiwan a significant relationship between the birth of babies who weigh less than 1500 grams and the prevalence of facial clefts (24). The maternal age of infants with clefts was mostly between 18 and 38 years which might be due to the higher frequency of the most the deliveries at this age range of mothers. Bille et al reported that older age of parents might have a role as a risk factor for cleft lip and palate (25).

## Conclusion

With the control of infectious disease and malnutrition, congenital anomalies including cleft lip and cleft palate are now making a proportionally greater contribution to the ill health in childhood in many countries. Our finding may be useful in identifying clues to the etiology of cleft lip and cleft palate. Once identified, the influencing factors may be prevented by proper intervention in the early stage of occurrence. The results may also have a role in planning and evaluating the strategies for primary prevention of cleft lip and cleft palate, particularly in high-risk populations.

## Data Availability

All data produced in the present study are available upon reasonable request to the authors

## Acknowledgments

We gratefully thank all the children and mothers in the region for their assistance in TRoCA programme over the past two decades. This study has been funded by the Tabriz University of Medical Sciences (IR.TBZMED.REC.1399.624).The authors report no conflict of interest.

